# A Predictive Model to Identify Complicated *Clostridiodes difficile* Infection

**DOI:** 10.1101/2022.05.18.22275113

**Authors:** Jeffrey A. Berinstein, Calen A. Steiner, Samara Rifkin, D. Alexander Perry, Dejan Micic, Daniel Shirley, Peter D.R. Higgins, Vincent B. Young, Allen Lee, Krishna Rao

## Abstract

**Background:** *Clostridioides difficile* infection (CDI) is a leading cause of healthcare-associated infections and may result in organ dysfunction, colectomy, and death. We recently showed that published risk scores to predict severe complications from CDI demonstrate poor performance upon external validation. We hypothesized that building and validating a model using geographically and temporally distinct cohorts would more accurately identify patients at risk for complicated CDI.

**Methods:** We conducted a multi-center retrospective cohort study of adult subjects diagnosed with CDI in the US. After randomly partitioning the data into training/validation set, we developed and compared three machine learning algorithms (Lasso regression, random forest, stacked ensemble models) with 10-fold cross-validation that used structured EHR data collected within 48 hours of CDI diagnosis to predict disease-related complications from CDI (intensive care unit admission, colectomy, or death attributable to CDI within 30 days of diagnosis). Model performance was assessed using area under the receiver operating curve (AUC).

**Results:** A total of 3,762 patients with CDI were included of which 218 (5.8%) had complications. Lasso regression, random forest, and stacked ensemble models all performed well with AUC ranging between 0.89-0.9. Variables of importance were similar across models, including albumin, bicarbonate, change in creatinine, systolic blood pressure, non-CDI-related ICU admission, and concomitant non-CDI antibiotics. Sensitivity analyses indicated that model performance was robust even when varying derivation cohort inclusion and CDI testing approach.

**Conclusion:** Using a large heterogeneous population of patients, we have developed and validated a prediction model based on structured EHR data that accurately estimates risk for complications from CDI.

**Key Points:** Machine learning models using structured electronic health records can be leveraged to accurately predict risk of severe complications related to *Clostridiodes difficile* infection, including intensive care unit admission, colectomy, and/or death.

## Introduction

*Clostridiodes* (*Clostridium) difficile* infection (CDI) is the leading cause of healthcare-associated infection in United States (U.S.) hospitals, accounting for nearly half a million infections per year [1,2]. Furthermore, up to 8% of patients with CDI may develop complicated disease, which is associated with worse outcomes, including organ dysfunction, severe sepsis, colectomy, and death [3]. In addition to the substantial patient morbidity and mortality, CDI has been attributed to $4.8 billion in acute healthcare costs in the United States, with even more costs associated with non-acute-care settings [4].

While multiple effective medical therapies have been developed, such as vancomycin, fidaxomicin, monoclonal antibodies, and fecal microbiota transplant [5], it remains unclear which patients are highest risk for a complicated disease course and therefore should receive more aggressive upfront therapy to improve outcomes. Current risk scores developed to identify patients at high risk for complicated CDI, have limited generalizability due to being developed in small, single-center cohorts or having not undergone rigorous external or prospective validation [5–14]. Recently, our group demonstrated that published CDI severity scoring systems performed poorly when tested on a large, multicenter cohort within the US [15]. Thus, an accurate prediction model that can be easily applied early after CDI diagnosis is needed to identify patients at risk for complicated CDI and to facilitate effective therapy that minimizes the risk for adverse CDI outcomes. In this study, we aimed to determine whether using structured electronic health record data from several geographically distinct centers in the US would provide a more generalizable predictive model for complicated CDI.

## Methods

### Patient Cohorts

We conducted a multi-center retrospective longitudinal cohort study at four geographically and temporally distinct cohorts, including the University of Michigan (2010-2012, 2015-2016), University of Wisconsin (2014-2015), and University of Chicago (2013-2015), as previously described by Perry *et al*. [15]. Adult subjects ≥18 years who were diagnosed with CDI were included in our analysis. CDI was diagnosed by presence of diarrhea (≥3 unformed stools in a 24-hour period) and positive real-time PCR for the *tcdB* gene (Simplexa *C. difficile* Universal Direct, Diasorin Molecular LLC, Cypress, CA) at University of Wisconsin and University of Chicago. Meanwhile, at both University of Michigan cohorts, CDI was diagnosed by presence of diarrhea (≥3 unformed stools in a 24-hour period) and a positive stool test for toxigenic *C. difficile* (positive testing for both the glutamate dehydrogenase [GDH] antigen and TcdA/TcdB by enzyme immunoassay [C. Diff Quik Chek Complete, Alere, Waltham, MA], or real-time polymerase chain reaction for the *tcdB* gene performed when GDH/toxin results were discordant). Patients with positive laboratory testing for CDI were identified using electronic health record (EHR).

### Primary Outcome

We defined CDI as complicated if it led to any of three adverse outcomes within 30 days of CDI diagnosis: admission to intensive care unit (ICU), colectomy, or death attributable to CDI as determined by the study team physicians [13,17]. Clinical and demographic variables including comorbidities, medications, vitals, laboratory results, and study results (such as radiographic imaging) were collected for each patient’s admission through automated query of the EHR. The institutional review boards at the University of Michigan, University of Chicago, and University of Wisconsin gave ethical approval for this work.

### Predictor Variables

A total of 32 predictor variables were evaluated, including demographic, biometric, biochemical, and co-morbid conditions (see Supplementary Table 1). Variable inclusion was based on literature review of clinically relevant factors. Non-CDI-related ICU admission and non-CDI concurrent antibiotics were included as predictor variables but only if ICU admission and/or antibiotic use were unrelated to CDI.

### Data Pre-Processing

We used R version 4.0.2 (R Foundation for Statistical Computing, Vienna, Austria) for cleaning and wrangling data. Data were randomly split with 75% of the data used for model training and the remaining 25% data held out for testing and validation. The data were stratified by the proportion of severe CDI events so that the distribution of the outcome was maintained in both the training and test set. Missing data were imputed using random forest-based imputation strategies (i.e., using the R package missForest) [18], which has previously been shown to have the lowest imputation error for both continuous and categorical variables [19]. Numeric variables were centered and scaled while categorical variables were recoded into dummy variables.

### Model Development and Testing

We developed three separate machine learning classification models using the *Tidymodels* framework [20], including L1-regularized logistic regression (least absolute shrinkage and selection operator or Lasso) using the R package *glmnet* [21], random forest with the R package *ranger* [22], and extreme gradient boosted trees with the R package *XGBoost* [23]. The best performing algorithms were combined in an ensemble model using stacking with the R package *stacks* [24]. Machine learning algorithms were first applied to training data to parameterize and fit the model. Ten-fold cross-validation was utilized to tune model hyperparameters. To evaluate the prediction accuracy of machine learning models, area under the receiver operating characteristic (ROC) curves (AUC) were calculated for each model using independent test data.

### Variable Importance

Variable importance was determined by permutation-feature importance analysis using the R package *vip* [25]. Permutation-feature importance measures the increase in prediction error when the variable is permuted and the relationship between variable and outcome is broken, thus the drop in the model score is indicative of how much the model depends on the feature [26]. To further improve interpretability of our machine learning models, we also performed locally interpretable model-agnostic explanations using the R package *breakDown*, which decomposes model predictions into parts that can be attributed to particular variables [27].

### Sensitivity Analyses

To determine whether our models were generalizable across sites and time, we performed a sensitivity analysis where we trained the machine learning algorithms on three cohorts and validated model performance on the fourth cohort. We repeated this process three times so that models were validated on each individual cohort.

Furthermore, as the definition of CDI varied by individual centers, we performed an additional sensitivity analysis to determine whether diagnosis of CDI by PCR only versus two-step mechanism influenced our model predictions. Sites that diagnosed CDI by PCR only (i.e. University of Wisconsin and University of Chicago) were analyzed separately from sites that utilized a two-step diagnostic approach (i.e. University of Michigan 2010 and 2016). We randomly split the data, training the model on 75% of the data and tested and validated our models on the remaining held-out 25% of the data. Model performance was evaluated as described above.

## Results

### Patient Characteristics

A total of 3,762 patients testing positive for *C. difficile* were collected from four cohorts at three sites from 01/01/2010 to 12/31/2015 (**Table 1)**. Overall, the mean age was 56.49 ± 19.82 years, 53.5% female, 68.6% Caucasian, 25.7% Black (with the highest proportion in the University of Chicago cohort), and 5.7% reported another race or not specified. A total of 218 patients (5.8%) met the primary endpoint, including 65 (4.5%) at the University of Chicago cohort; 90 (7.9%) at the University of Michigan 2010-2012 cohort; 28 subjects (4.4%) at the University of Michigan 2010-2012 cohort; and 35 (6.5%) at the University of Wisconsin.

**Table 1:**
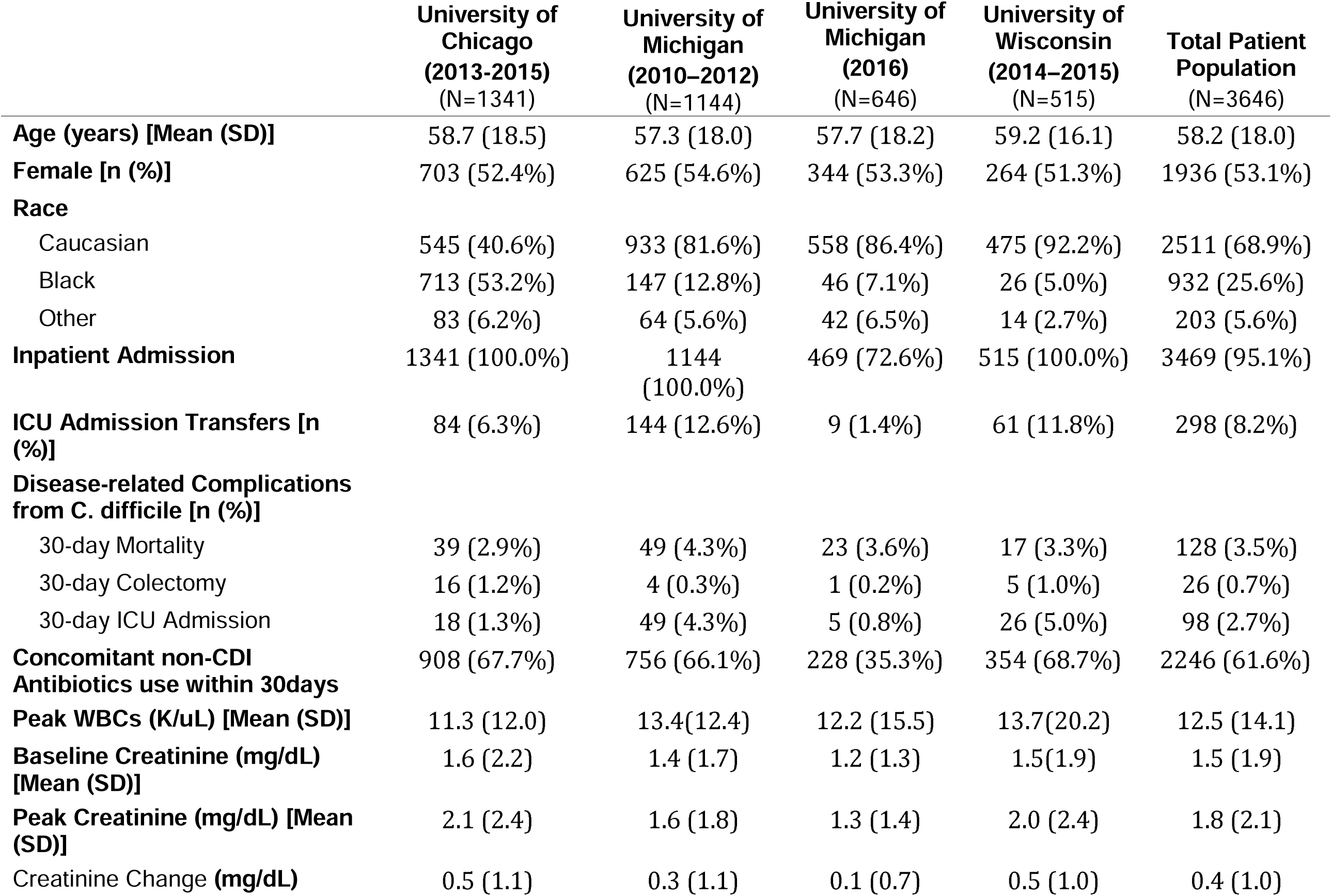
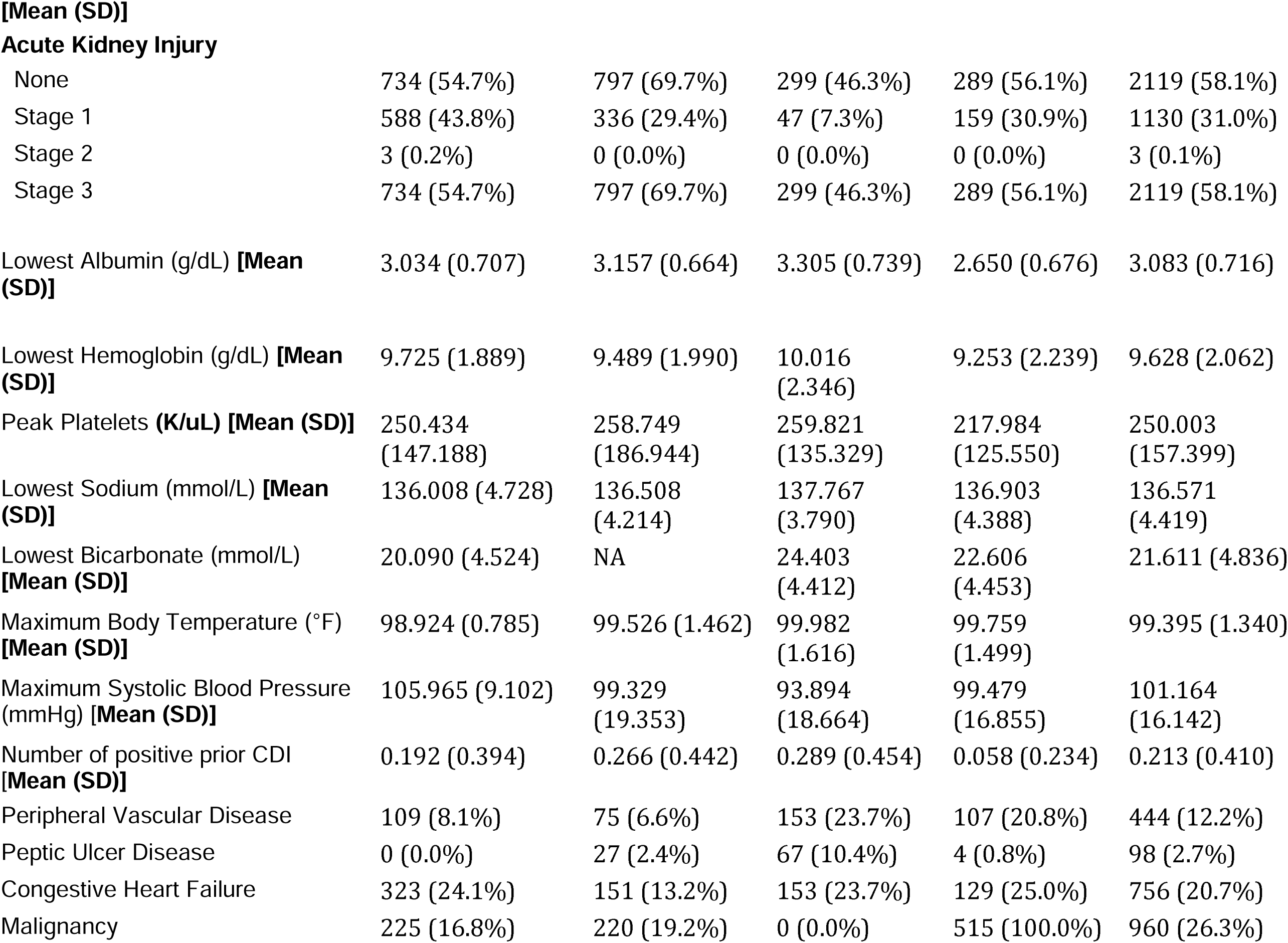
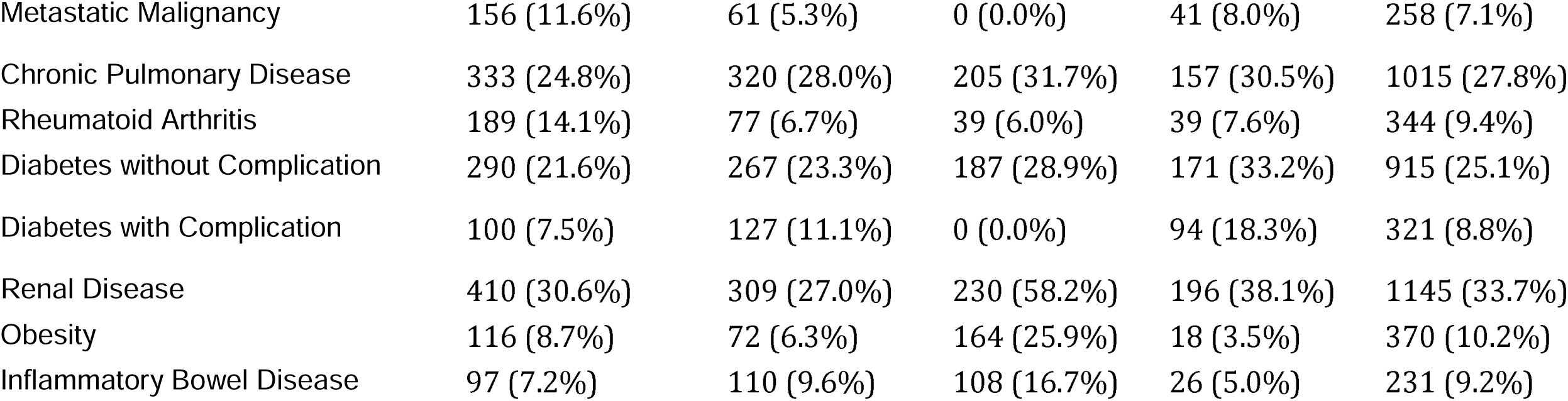
Patient Characteristics.

|  | University of<br>Chicago<br>(2013-2015)<br>(N=1341) | University of<br>Michigan<br>(2010–2012)<br>(N=1144) | University of<br>Michigan<br>(2016)<br>(N=646) | University of<br>Wisconsin<br>(2014–2015)<br>(N=515) | Total Patient<br>Population<br>(N=3646) |
| --- | --- | --- | --- | --- | --- |
| **Age (years) [Mean (SD)]** | 58.7 (18.5) | 57.3 (18.0) | 57.7 (18.2) | 59.2 (16.1) | 58.2 (18.0) |
| **Female [n (%)]** | 703 (52.4%) | 625 (54.6%) | 344 (53.3%) | 264 (51.3%) | 1936 (53.1%) |
| **Race** |  |  |  |  |  |
| Caucasian | 545 (40.6%) | 933 (81.6%) | 558 (86.4%) | 475 (92.2%) | 2511 (68.9%) |
| Black | 713 (53.2%) | 147 (12.8%) | 46 (7.1%) | 26 (5.0%) | 932 (25.6%) |
| Other | 83 (6.2%) | 64 (5.6%) | 42 (6.5%) | 14 (2.7%) | 203 (5.6%) |
| **Inpatient Admission** | 1341 (100.0%) | 1144<br>(100.0%) | 469 (72.6%) | 515 (100.0%) | 3469 (95.1%) |
| **ICU Admission Transfers [n**<br>**(%)]** | 84 (6.3%) | 144 (12.6%) | 9 (1.4%) | 61 (11.8%) | 298 (8.2%) |
| **Disease-related Complications**<br>**from C. difficile [n (%)]** |  |  |  |  |  |
| 30-day Mortality | 39 (2.9%) | 49 (4.3%) | 23 (3.6%) | 17 (3.3%) | 128 (3.5%) |
| 30-day Colectomy | 16 (1.2%) | 4 (0.3%) | 1 (0.2%) | 5 (1.0%) | 26 (0.7%) |
| 30-day ICU Admission | 18 (1.3%) | 49 (4.3%) | 5 (0.8%) | 26 (5.0%) | 98 (2.7%) |
| **Concomitant non-CDI**<br>**Antibiotics use within 30days** | 908 (67.7%) | 756 (66.1%) | 228 (35.3%) | 354 (68.7%) | 2246 (61.6%) |
| **Peak WBCs (K/uL) [Mean (SD)]** | 11.3 (12.0) | 13.4(12.4) | 12.2 (15.5) | 13.7(20.2) | 12.5 (14.1) |
| **Baseline Creatinine (mg/dL)**<br>**[Mean (SD)]** | 1.6 (2.2) | 1.4 (1.7) | 1.2 (1.3) | 1.5(1.9) | 1.5 (1.9) |
| **Peak Creatinine (mg/dL) [Mean**<br>**(SD)]** | 2.1 (2.4) | 1.6 (1.8) | 1.3 (1.4) | 2.0 (2.4) | 1.8 (2.1) |
| **Creatinine Change (mg/dL)** | 0.5 (1.1) | 0.3 (1.1) | 0.1 (0.7) | 0.5 (1.0) | 0.4 (1.0) |
| **Acute Kidney Injury** |  |  |  |  |  |
| None | 734 (54.7%) | 797 (69.7%) | 299 (46.3%) | 289 (56.1%) | 2119 (58.1%) |
| Stage 1 | 588 (43.8%) | 336 (29.4%) | 47 (7.3%) | 159 (30.9%) | 1130 (31.0%) |
| Stage 2 | 3 (0.2%) | 0 (0.0%) | 0 (0.0%) | 0 (0.0%) | 3 (0.1%) |
| Stage 3 | 734 (54.7%) | 797 (69.7%) | 299 (46.3%) | 289 (56.1%) | 2119 (58.1%) |
| Lowest Albumin (g/dL) **[Mean (SD)]** | 3.034 (0.707) | 3.157 (0.664) | 3.305 (0.739) | 2.650 (0.676) | 3.083 (0.716) |
| Lowest Hemoglobin (g/dL) **[Mean (SD)]** | 9.725 (1.889) | 9.489 (1.990) | 10.016 (2.346) | 9.253 (2.239) | 9.628 (2.062) |
| Peak Platelets (K/uL) **[Mean (SD)]** | 250.434 (147.188) | 258.749 (186.944) | 259.821 (135.329) | 217.984 (125.550) | 250.003 (157.399) |
| Lowest Sodium (mmol/L) **[Mean (SD)]** | 136.008 (4.728) | 136.508 (4.214) | 137.767 (3.790) | 136.903 (4.388) | 136.571 (4.419) |
| Lowest Bicarbonate (mmol/L) **[Mean (SD)]** | 20.090 (4.524) | NA | 24.403 (4.412) | 22.606 (4.453) | 21.611 (4.836) |
| Maximum Body Temperature (°F) **[Mean (SD)]** | 98.924 (0.785) | 99.526 (1.462) | 99.982 (1.616) | 99.759 (1.499) | 99.395 (1.340) |
| Maximum Systolic Blood Pressure (mmHg) **[Mean (SD)]** | 105.965 (9.102) | 99.329 (19.353) | 93.894 (18.664) | 99.479 (16.855) | 101.164 (16.142) |
| Number of positive prior CDI **[Mean (SD)]** | 0.192 (0.394) | 0.266 (0.442) | 0.289 (0.454) | 0.058 (0.234) | 0.213 (0.410) |
| Peripheral Vascular Disease | 109 (8.1%) | 75 (6.6%) | 153 (23.7%) | 107 (20.8%) | 444 (12.2%) |
| Peptic Ulcer Disease | 0 (0.0%) | 27 (2.4%) | 67 (10.4%) | 4 (0.8%) | 98 (2.7%) |
| Congestive Heart Failure | 323 (24.1%) | 151 (13.2%) | 153 (23.7%) | 129 (25.0%) | 756 (20.7%) |
| Malignancy | 225 (16.8%) | 220 (19.2%) | 0 (0.0%) | 515 (100.0%) | 960 (26.3%) |
| Metastatic Malignancy | 156 (11.6%) | 61 (5.3%) | 0 (0.0%) | 41 (8.0%) | 258 (7.1%) |
| Chronic Pulmonary Disease | 333 (24.8%) | 320 (28.0%) | 205 (31.7%) | 157 (30.5%) | 1015 (27.8%) |
| Rheumatoid Arthritis | 189 (14.1%) | 77 (6.7%) | 39 (6.0%) | 39 (7.6%) | 344 (9.4%) |
| Diabetes without Complication | 290 (21.6%) | 267 (23.3%) | 187 (28.9%) | 171 (33.2%) | 915 (25.1%) |
| Diabetes with Complication | 100 (7.5%) | 127 (11.1%) | 0 (0.0%) | 94 (18.3%) | 321 (8.8%) |
| Renal Disease | 410 (30.6%) | 309 (27.0%) | 230 (58.2%) | 196 (38.1%) | 1145 (33.7%) |
| Obesity | 116 (8.7%) | 72 (6.3%) | 164 (25.9%) | 18 (3.5%) | 370 (10.2%) |
| Inflammatory Bowel Disease | 97 (7.2%) | 110 (9.6%) | 108 (16.7%) | 26 (5.0%) | 231 (9.2%) |

### Model Training and Performance

Data were randomly split into a training set of 2,763 cases and an independent validation set of 921 cases. Splitting of the data set was stratified by the proportion of severe CDI to preserve the distribution of complicated CDI in both the training and validation set. We found that lasso regression, random forest, and stacked ensemble models all performed well with AUC scores ranging from 0.88-0.89 (**Figure 1**) when tested on an independent test dataset. However, XGBoost models showed poor performance (AUC 0.50) and were not carried forward in subsequent analyses (data not shown). The random forest model performed marginally better than the lasso regression and stacked ensemble model, but only with a magnitude of 0.01, which is unlikely to be clinically significant. Model calibration plots for the three models are illustrated in **Supplemental Figure 1**.

**Figure 1.**
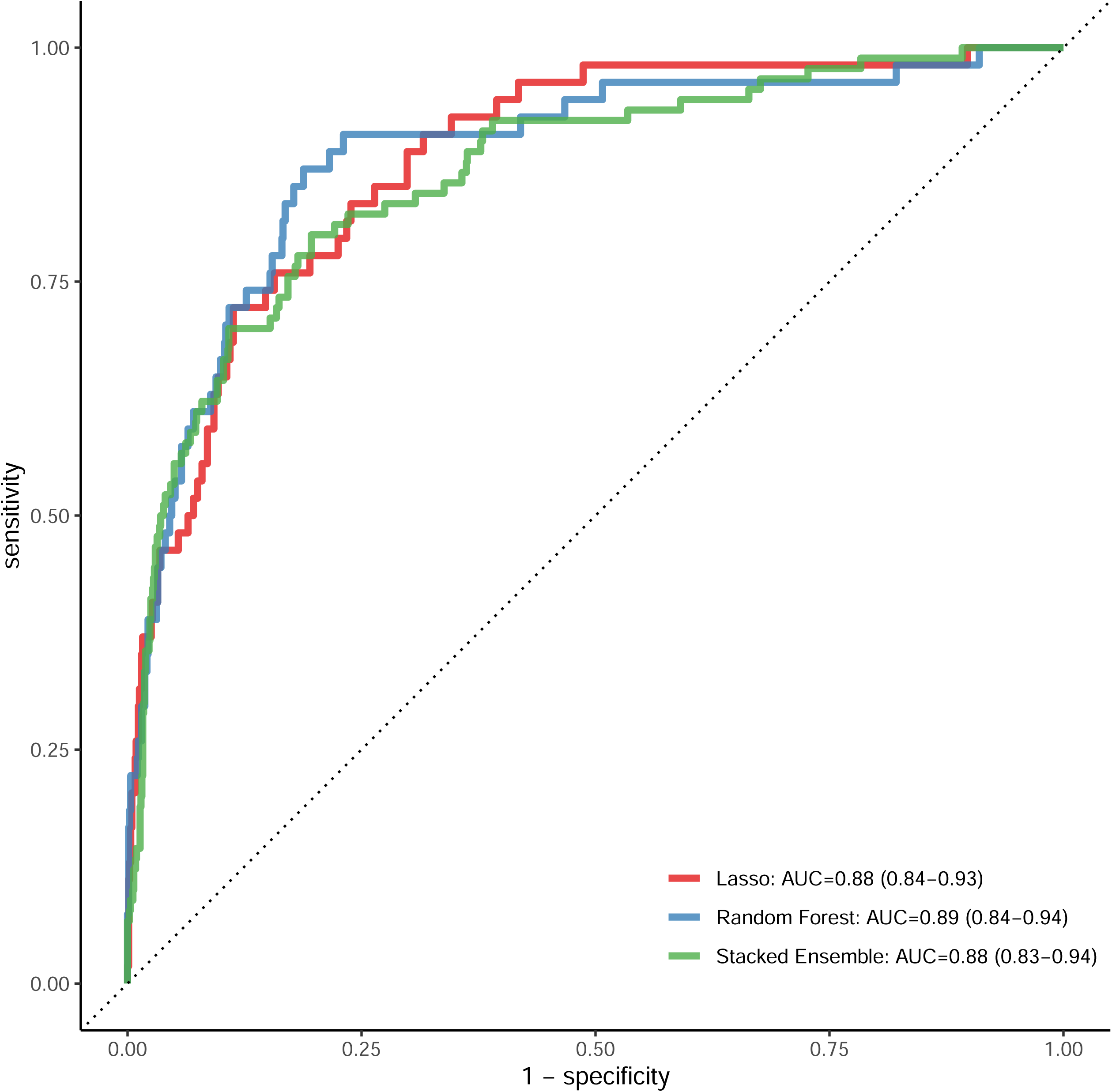
Receiver operator characteristic (ROC) curves for Lasso, Random Forest, and Stacked Ensemble models. After randomly splitting the data into training/validation sets, Lasso regression (*red line*), Random Forest (*blue line*), and Stacked Ensemble (*green line*) models were trained. All models demonstrated excellent performance when tested on an independent validation set (Lasso regression: AUC = 0.88 [95% CI 0.84-0.93]; Random Forest: AUC = 0.89 [95% CI 0.84-0.94]; Stacked Ensemble: AUC = 0.88 [95% CI 0.83-0.94]).

#### Sensitivity Analyses

To test the generalizability of our multi-cohort approach as well as to assess for temporal trends and variations in management practices and participant composition among the various cohorts, we performed a sensitivity analysis by deriving models using only three of the four cohorts in the dataset and holding out the fourth cohort as the independent test set. We found that even models trained on only three cohorts were able to predict severe CDI in the fourth cohort left out of the training set with high accuracy. Model performance was robust in this sensitivity analysis (AUC ranging from 0.84-0.92) although there was a drop in performance when data from the University of Chicago was held out and used as the test set (AUC 0.75-0.76) **(Figure 2**).

**Figure 2.**
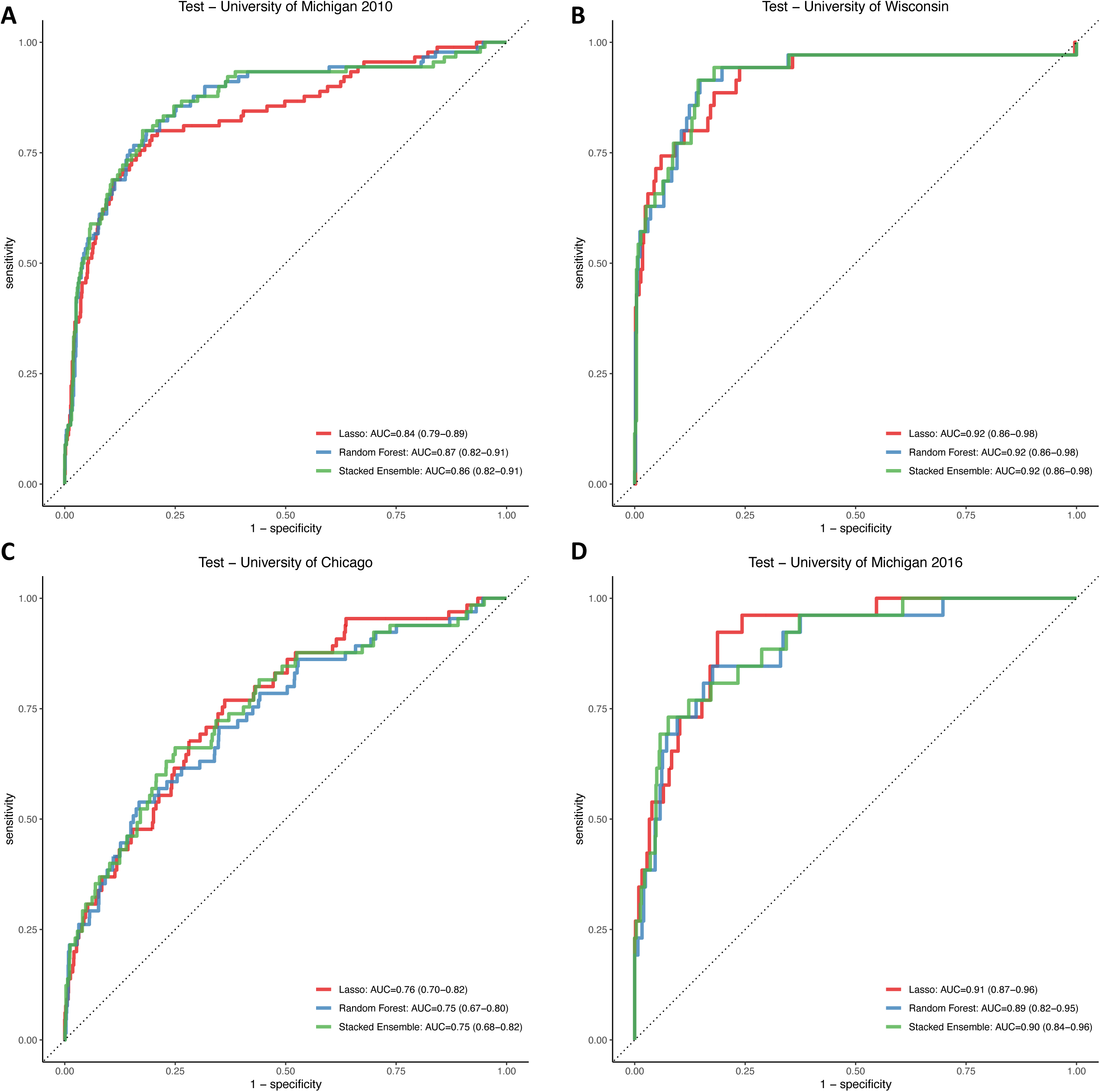
Models are robust despite geographical, demographic, and temporal variability. To determine whether models were generalizable across centers and across time, machine learning algorithms were derived and trained using data from three cohorts and then validated model performance on the fourth cohort (labeled as Test in the figure). This process was repeated three times so that models were validated on each individual cohort. The Lasso regression (*red line*), Random Forest (*blue line*), and Stacked ensemble (*green line*) models demonstrated good performance when tested on (**A**) the University of Michigan 2010-2012 cohort (Lasso regression: AUC = 0.84 [95% CI 0.79-0.89); Random Forest: AUC = 0.87 [95% CI 0.82-0.91]; Stacked Ensemble: AUC = 0.86 [95% CI 0.82-0.91]); (**B**) University of Wisconsin cohort (Lasso regression: AUC = 0.92 [95% CI 0.86-0.98]; Random Forest: AUC = 0.92 [95% CI 0.86-0.98]; Stacked Ensemble: AUC = 0.92 [95% CI 0.86-0.98]); and (**D**) University of Michigan 2015-2016 cohort (Lasso regression: AUC = 0.91 [95% CI 0.87-0.96]; Random Forest: AUC = 0.89 [95% CI 0.82-0.95]; Stacked Ensemble: AUC = 0.90 [95% CI 0.84-0.96]). Performance of the models dropped but still showed adequate performance when tested on (**C**) the University of Chicago cohort (Lasso regression: AUC = 0.76 [95% CI 0.70-0.82]; Random Forest: AUC = 0.75 [95% CI 0.67-0.80]; Stacked Ensemble: AUC = 0.75 [95% CI 0.68-0.82]).

In addition, to determine whether the method for CDI diagnosis may have impacted model performance, we performed separate sensitivity analyses using data from sites that used PCR alone (i.e. University of Chicago and University of Wisconsin) vs. two-step testing (i.e. University of Michigan 2010 and 2016) to train and validate models. For sites that used two-step testing, our models retained excellent performance (AUC 0.89 – 0.91), while model performance was lower when using sites that used PCR testing alone as the hold-out-test set (AUC 0.79 – 0.84) (**Supplemental Figure 2**).

### Predictive Features for Complicated CDI

We performed a permutation-based variable importance analysis to determine the variable importance of each predictor of complicated CDI (**Figure 3**). Interestingly, the variables of importance were similar across models, but varied in their relative contribution to each model. The top predictors shared across all models included albumin, bicarbonate, change in creatinine, systolic blood pressure, non-CDI-related ICU admission within 30 days, and concomitant non-CDI antibiotics within 30 days of CDI diagnosis. Model performance was similar despite differences in relative contributions by each variable.

**Figure 3.**
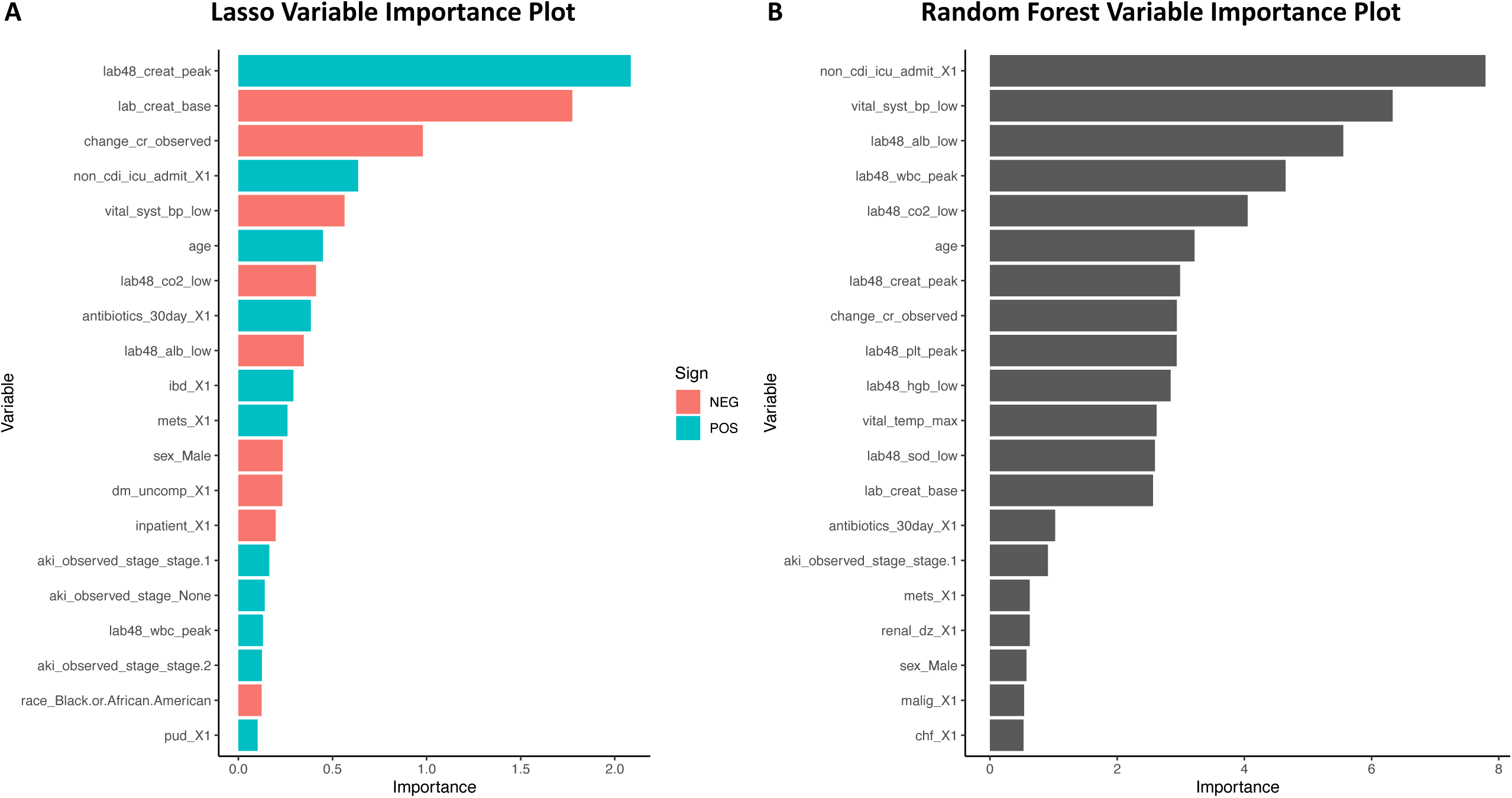
Variable Importance Plot for Lasso Regression and Random Forest. Twenty of the most important variables by permutation-based variable importance analysis are illustrated for (**A**) Lasso Regression and (**B**) Random Forest. Variables positively associated with complicated *Clostridiodes difficile* infection (CDI) are shown in *blue* while variables negatively associated with complicated CDI are shown in *orange*. Please note, determining the directionality of association for variables using permutation-based variable importance analysis with nonlinear based machine learning techniques, such as random forest, is not possible.

To enhance the interpretability of our machine learning models, we also performed breakdown plots to determine how each variable contributes to a final prediction. For patients with low predicted probability of complicated CDI (**Figure 4A-B**), absence of factors, such as non-CDI-related ICU admission, concurrent non-CDI antibiotics, low CO_2_ levels, low systolic blood pressure, and peak WBC count were associated with decreased risk for complicated CDI. In contrast, for patients with high predicted probability for complicated CDI (**Figure 4C-D**), non-CDI-related ICU admission, high WBC count, low systolic blood pressure, low albumin level, and concurrent non-CDI antibiotics were associated with increased risk for complicated CDI.

**Figure 4.**
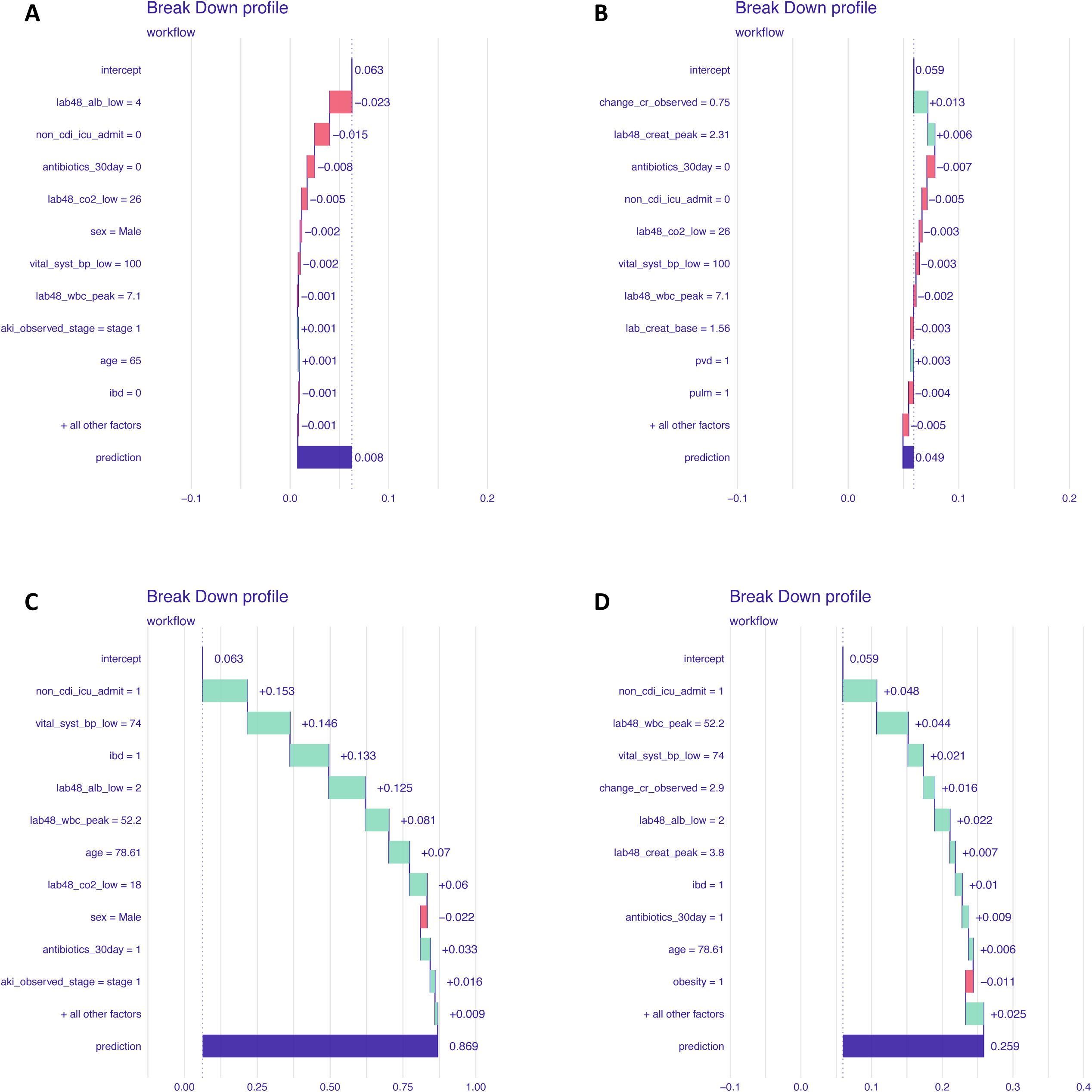
Breakdown plot for patients with low and high predicted probability for complicated *Clostridiodes difficile* infection (CDI). Breakdown plot is shown for (**A**) Lasso and (**B**) random forest model in the same patient (patient #1) with low predicted probability for CDI. Similarly, breakdown plot is depicted for (**C**) Lasso and (**D**) random forest model in a patient with high predicted probability for complicated CDI (patient #277). Intercept represents the mean model-specific predicted probability for complicated CDI while each subsequent variable increases or decreases predicted probability and results in the overall predicted probability (labeled prediction).

## Discussion

In this multi-site cohort study, machine learning models based on structured electronic health record data accurately predicted disease-related complications from CDI. All three machine learning methods, including lasso regression, random forest, and a stacked ensemble method demonstrated excellent performance in predicting severe complications from CDI (i.e., AUC of 88-89%). Importantly, we intentionally developed models without site-specific variables, and our results suggest that this model is generalizable across centers and time, which is critical when considering the heterogeneity in patient population and practice patterns across the United States. Furthermore, our results were generally agnostic to specific algorithms and performed equally well when using both linear and nonlinear machine learning approaches. Finally, although there was some model-specific variability, the predictors most important in discriminating severe complications attributable to CDI were similar between models.

While several CDI severity and complication scoring systems have been developed previously, they generally were developed from single centers and were not externally validated [5–14]. Our group recently published the largest external validation of published CDI severity scoring systems and found that all models yielded AUC scores below 70% suggesting that current models are not generalizable and cannot reliably predict severe complications from CDI [15].

There are multiple notable strengths to our analysis which make our approach generalizable. First, our models were developed using data from three geographically distinct sites, which were composed of a heterogenous population. When we validated our model on each site as an independent hold-out data set, our models continue to demonstrate good performance. Secondly, our models were robust despite differences in temporal trends and practice patterns across sites. For example, CDI was diagnosed by positive PCR test alone at the University of Chicago and the University of Wisconsin, while a two-step algorithm was required for positive CDI diagnosis at the University of Michigan. Furthermore, the University of Michigan started to use Vancomycin as first-line treatment for CDI starting in 2013 while the other centers did not switch from metronidazole to vancomycin as first-line treatment until mid-2016. Despite these differences in practice patterns and temporal trends, our sensitivity analyses demonstrated that our model retained good performance when deriving models from the three earlier cohorts and validating it on the latter University of Michigan cohort as well as deriving and validating models on the two sites that used PCR testing alone for CDI diagnosis compared to sites that used a two-step algorithm.

While our model showed strong performance overall, we did see a noticeable drop in performance on sensitivity analyses when the model was validated on the cohort from the University of Chicago. We speculate that this may be related to the diversity of the patients comprising the University of Chicago cohort that was not captured when this site was not included in deriving the model. Black individuals represented a larger proportion of the cohort at the University of Chicago compared with other sites, while Black race was associated with a higher likelihood of severe CDI outcomes. Thus, important information related to racial diversity may have been lost when model derivation was performed using only the University of Michigan and University of Wisconsin cohorts which were comprised of a small proportion of Black patients. We believe this also strengthens the rationale for building and validating our primary model using four geographically and temporally heterogeneous cohorts, which was critical in developing a more generalizable model for predicting complicated CDI.

We achieved similar classification performance when using lasso regression, random forest or combining these algorithms using a stacked ensemble method. While random forest models showed a numerically higher performance compared to the other models, the incremental benefit was likely negligible. Machine learning algorithms, such as random forest and stacked ensemble methods, do not produce coefficients and are inherently less interpretable compared with lasso regression [26]. Thus, transforming model results into a risk score is more complex. Given the ease of interpretation as well as reduced computational costs associated with regression-based approaches, our lasso regression model may be favored for future applications.

In general, variables that carried the greatest importance were consistent across models and across sites. Most variables identified as important predictors of complicated CDI by our model are also consistent with our clinical and biological understanding of CDI pathogenesis and progression, such as peak creatinine, peak white blood cells, low albumin levels, low blood pressure, low hemoglobin, advanced age, concurrent antibiotic use for treating non-CDI infections, low bicarbonate, and ICU admissions. Importantly, all variables were collected within 48 hours of CDI diagnosis, which increases the clinical utility of our model, by allowing for early calculation of patients at risk for developing complicated CDI. Leukocytosis (white blood cell count > 15,000 cells/mL) and acute rise in serum creatinine > 1.5 mg/dL are well established markers for severe CDI and were initially incorporated into guidelines by Society for Healthcare Epidemiology of America and the Infectious Disease Society of America in 2018 to determine which patients at risk for severe disease and should be offered more aggressive upfront therapy (i.e., vancomycin over metronidazole) [28]. In addition, older age [29,30], hypoalbuminemia [31], low hemoglobin [32], concurrent antibiotic use [31], and ICU admission [14,31] have also been previously identified as predictors of poor outcomes from CDI. Although ICU admission was part of our composite endpoint, we were careful to include only patients who were admitted to the ICU prior to CDI diagnosis as our predictor variable.

Our study has several notable strengths. To our knowledge, this is the largest study combining data from four distinct cohorts composed of both temporally and geographically distinct patients to create a predictive model of complicated CDI. This significantly improved the generalizability of the model. Previous models predicting severe or complicated CDI were mostly derived from smaller single centers. By using rigorous and well-validated predictive modeling techniques and by comparing several different predictive modeling approaches, we were able to develop a highly accurate model for predicting complicated CDI using readily available structured EHR data. In addition, the use of permutation-based variable importance analysis allowed us to identify the importance of each predictor in our models. By improving the interpretability of our model, we anticipate this will enhance clinical utility, provider buy-in, and uptake of use when implemented.

These results should be interpreted in the context of several limitations. First, our analysis is retrospective. Prospective studies are necessary for understanding how such models perform in real time. Second, our models were derived from four cohorts from major academic medical centers in the midwestern United States. Model performance will have to be evaluated outside of this setting to confirm generalizability as patient populations, clinical protocols, and risk factors may vary across institutions. Third, given that our model merely identified associations, additional prospective investigation is required to establish the direction of the true underlying relationships (e.g., through RCTs). Moreover, despite the removal of variables that could serve as proxies for our composite outcome, some model features may not be true risk factors but rather markers for the beginning of complicated CDI itself. In the future, this may be elucidated by tracking the evolution of a patient’s risk over multiple days of their hospitalization. Fourth, we cannot exclude the possibility that patients may experience the outcome at another hospital, and thus we may have potentially underestimated the extent of complications from CDI. In addition, although CDI testing was recommended only for symptomatic patients during our study period and this was further validated by chart review, some positive CDI tests might reflect asymptomatic carriers. Lastly, our model employed a large number of variables which may affect clinician-perceived useability; however, this would be mitigated by embedding a decision tool which utilizing our prediction models directly within the EHR.

In summary, this multi-center cohort study comprised of a large heterogeneous population demonstrates that machine learning algorithms based on structured EHR data can accurately estimate patients’ risk for developing complications from CDI. Our approach leverages variables that can be readily extracted from the EHR early in the course of a patient’s CDI. This approach has many potential applications for guiding clinician decision making in the management of CDI. Future studies may determine whether prospective deployment of this model may aid clinicians to tailor patient therapy in real-time and allow for early use of more aggressive therapies to minimize risk of complications.

## Data Availability

All data produced in the present study are available upon reasonable request to the authors

## Abbreviations

AUC: area under the receiver operator characteristic curve
CDI: *Clostridiodes difficile* infection
EHR: electronic health records
ICU: intensive care unit

## Data Availability

All data produced in the present study are available upon reasonable request to the authors

## Figure Legends

**Supplemental Figure 1**. Calibration plot for Lasso, Random Forest, and Stacked Ensemble Models. The calibration plots fitted with a locally estimated scatterplot smoothing (loess) curve for the Lasso regression (*red line*), Random Forest (*blue line*), and Stacked Ensemble (*green line*) models show overall good agreement between observed and predicted outcomes. However, the Lasso regression and Random Forest models tend to overestimate risk for complicated CDI at low probabilities while the Stacked Ensemble model underestimates the risk for complicated CDI at low probabilities.

**Supplemental Figure 2**. Models retain good performance despite differences in definitions for *Clostridiodes difficile* infection (CDI) across centers. As the definition for CDI employed at the University of Chicago and the University of Wisconsin differed from that used at the University of Michigan, we performed a sensitivity analysis to see if our model was robust to these differences. (**A**) Using data only from the University of Chicago and University of Wisconsin cohorts which were randomly split 75/25 into training/validation sets, machine learning algorithms were trained and then validated on the independent test set. Lasso models (*red line*), random forest (*blue line*), and stacked ensemble (*green line*) showed good performance when validated on the independent test set. (**B**) This process was repeated but using data only from the University of Michigan 2010 and 2016 cohorts. From University of Chicago and University of Wisconsin and then validated model performance on both University of Michigan cohorts (labeled as Michigan 2010 and Michigan 2016).

