## Supplementary figures and images for "A Predictive Model to Identify Complicated *Clostridiodes difficile* Infection"

### Supplemental Figure 1

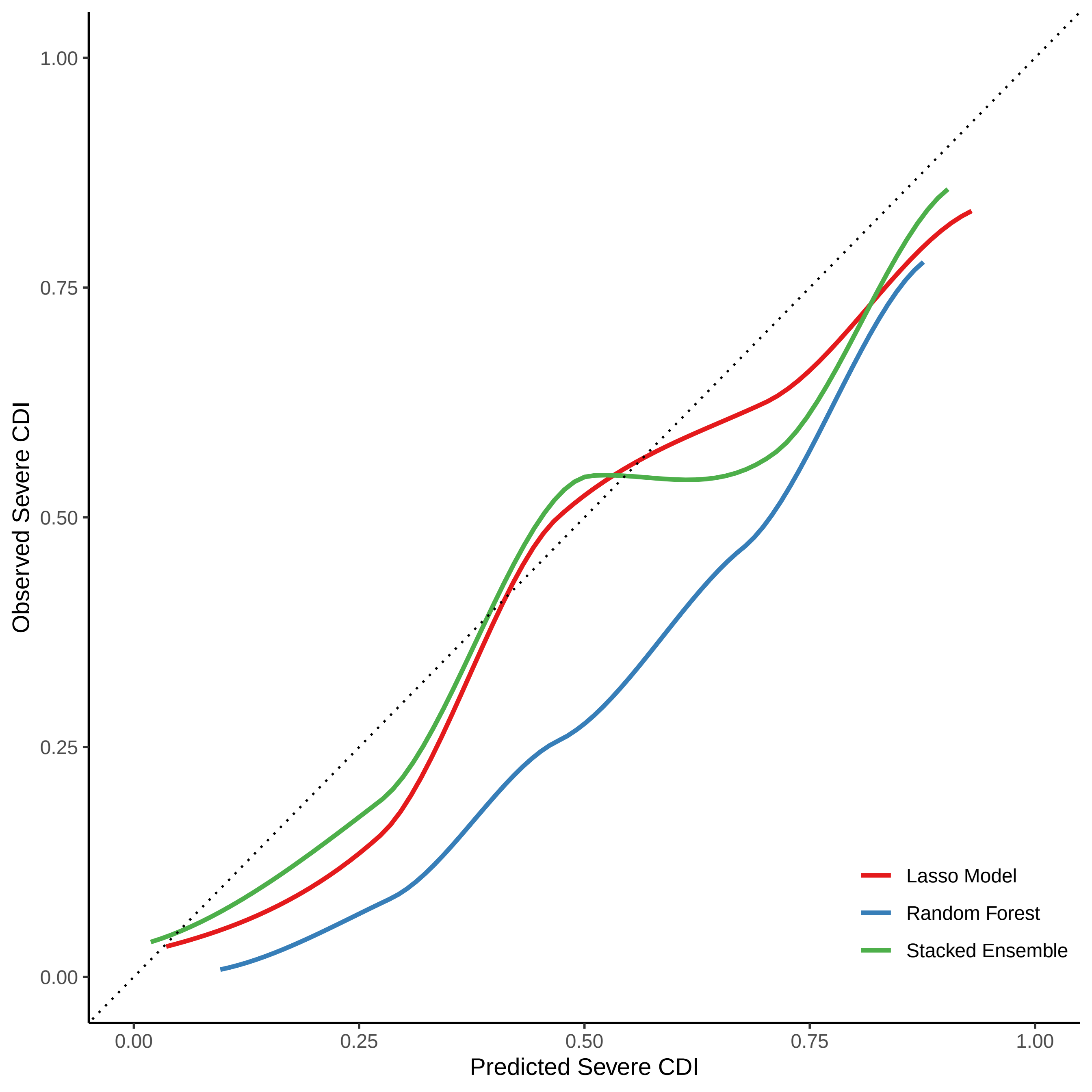

### Supplemental Figure 2

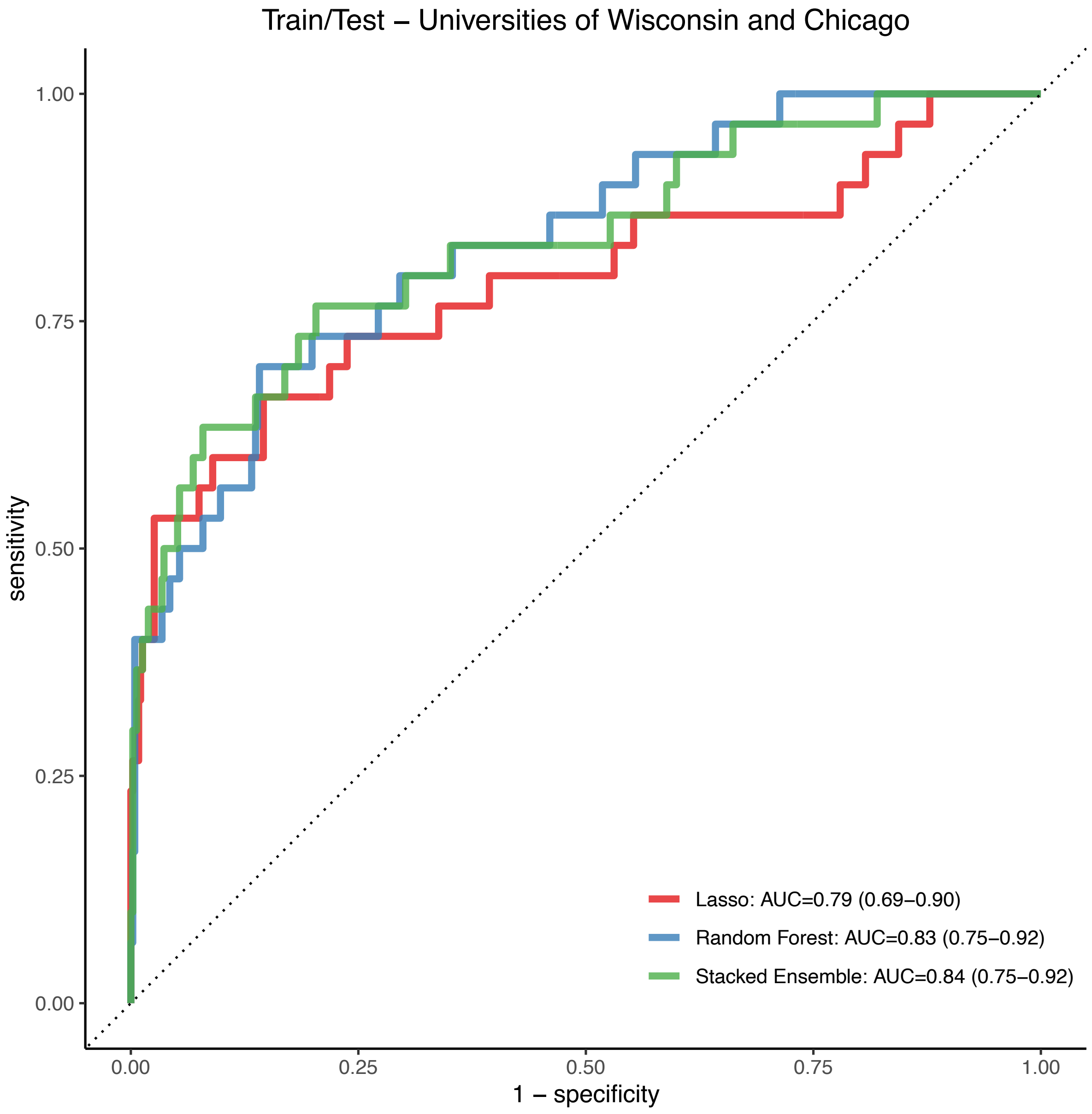

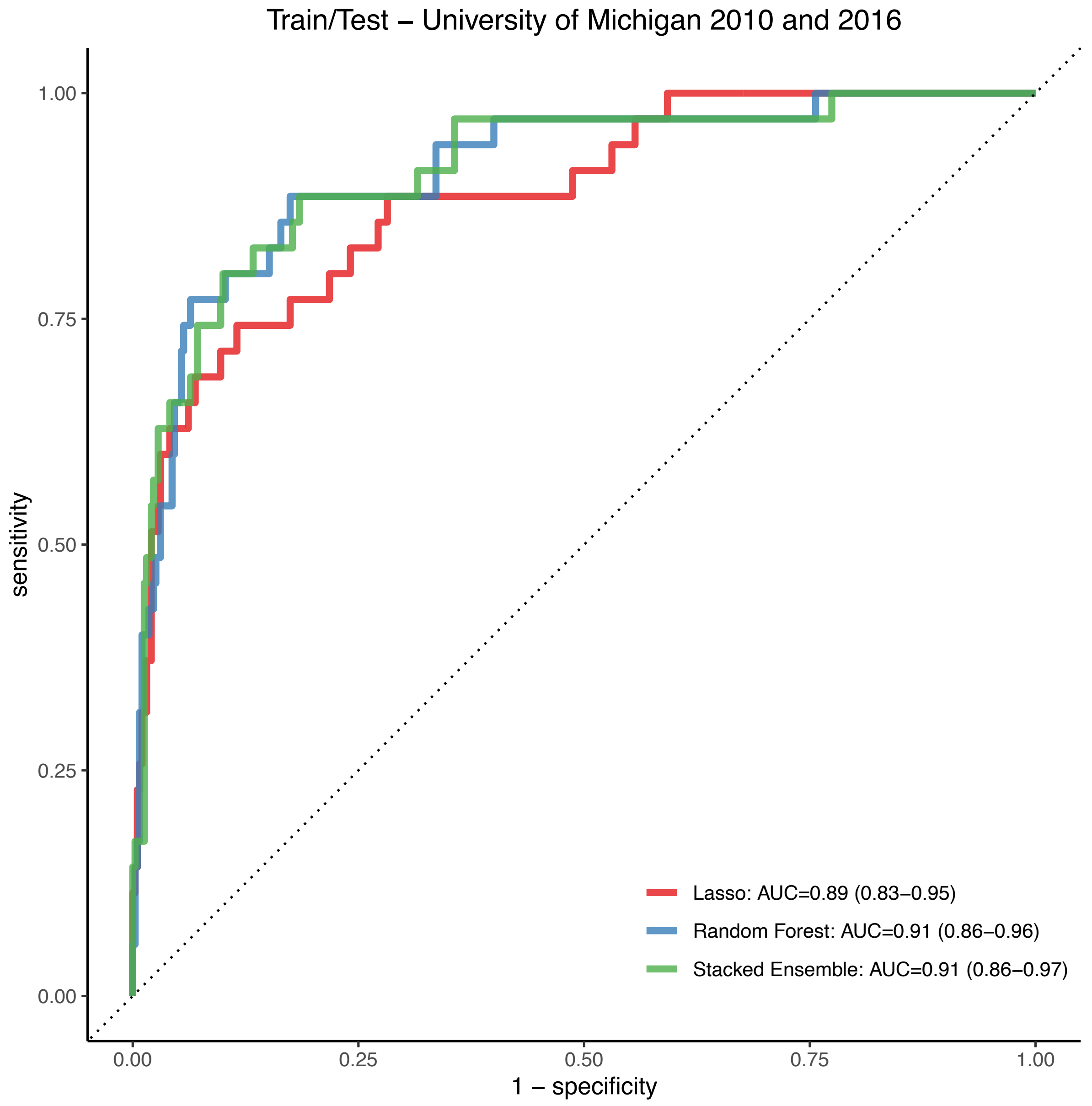
