## Supplemental Table 1 for "A Predictive Model to Identify Complicated *Clostridiodes difficile* Infection"

**Supplemental Table 1: Variables Evaluated**

| **Category** | **Variable** |
| --- | --- |
| Demographics | Age, sex, race/ethnicity |
| Labs | Baseline creatinine, peak Creatinine, nadir albumin, low hemoglobin, peak platelet, nadir bicarbonate, peak white blood cell, nadir sodium, change in creatinine |
| Vital Signs | Peak temperature, nadir systolic blood pressure |
| Hospital Course | Inpatient admission, acute kidney injury, non-CDI-related antibiotics within 30 days, ventilator status, non-CDI-related intensive care unit admission |
| Comorbidities | Inflammatory bowel disease, chronic kidney disease, obesity, malignancy, malignancy with metastasis, diabetes mellitus, diabetes mellitus with complications, rheumatologic disease, peptic ulcer disease, peripheral vascular disease, pulmonary disease, congestive heart failure, |
